# QUADRICEPS STRENGTH AND KNEE ABDUCTION MOMENT DURING LANDING IN ADOLESCENT ATHLETES

**DOI:** 10.64898/2026.03.06.26347192

**Authors:** Lillian Johnson, Colin W. Bond, Benjamin C. Noonan

**Author notes:** **Corresponding Author:** Colin W. Bond, PhD, MBA, Sports Science and Biomedical Engineering Laboratory, 1711 South University Dr., Fargo, ND 58103. **Ethical Statement:** The Sanford Health Institutional Review Board gave ethical approval for this work (Study ID: 2976). **Data Availability:** Data produced in the present study are available upon reasonable request to the authors. **Funding:** This study did not receive any funding.

## Abstract

**Background:** Quadriceps weakness may reduce sagittal plane shock absorption during landing, shifting load toward the frontal plane and increasing knee abduction moment (KAM), a biomechanical risk factor for anterior cruciate ligament (ACL) injuries.

**Purpose:** The purpose of this study was to evaluate the association between isokinetic quadriceps strength and peak KAM during drop vertical jump landing in adolescent athletes.

**Study Design:** Secondary analysis of previously collected data.

**Methods:** Healthy adolescent athletes completed quadriceps strength testing using an isokinetic dynamometer and a biomechanical assessment during a drop vertical jump task. Quadriceps strength was quantified as peak concentric torque and the peak external KAM was calculated during the landing phase on the dominant limb. Both strength and KAM were normalized to body mass. Linear regression was used to examine the association between normalized quadriceps strength and peak external KAM on the dominant limb.

**Results:** The association between quadriceps strength and peak normalized KAM on the dominant limb was not statistically significant (β = −0.053 (95% CI [−0.137 to 0.030]), F(1,119) = 1.62, R^2^ = 0.013, p = 0.206). Quadriceps strength explained only 1.3% of the variance in peak KAM, indicating a negligible association between these variables in this cohort.

**Discussion:** Quadriceps strength was not associated with peak normalized KAM during landing, suggesting that frontal-plane knee loading during a drop vertical jump is not meaningfully explained by maximal concentric quadriceps strength alone. KAM appears to be driven more by multi-joint movement strategy and neuromuscular coordination than by the capacity of a single muscle group.

## INTRODUCTION

The influence of quadriceps strength on compensatory movement patterns—specifically, the peak knee abduction moment (KAM), which often occurs early after landing from a jump—in the context of anterior cruciate ligament (ACL) injury risk is not well understood. The ACL is a key stabilizer of the knee joint, and injuries to this structure are especially common among athletes who perform frequent cutting, pivoting, and deceleration movements.^1,2^ These high-demand activities place significant stress on the lower extremity, requiring it to attenuate these loads.^1,2^ Most ACL injuries arise from non-contact mechanisms, sparking extensive research into the biomechanical risk factors associated with this clinically and economically burdensome injury.^3^ One area that requires further investigation is the role of quadriceps muscle weakness and compensatory high-risk force attenuation strategies in the frontal plane at the knee joint. Understanding this relationship may enhance the sensitivity of ACL injury risk screening and guide targeted prevention strategies for individuals who demonstrate suboptimal movement patterns.

Controlled knee flexion—primarily driven by the large, phasic quadriceps—is the optimal mechanism for force attenuation used by athletes during cutting, pivoting, and deceleration movements. This spring-like behavior of the knee makes sagittal plane motion both biomechanically efficient and protective against injury.^3,4^ However, athletes may adopt a suboptimal motor control strategy that could shift load attenuation into the frontal plane, manifesting in part as an elevated KAM.^1,5^

An elevated KAM, which results from increased and misaligned ground reaction forces (GRFs), has emerged as a well-established biomechanical predictor of ACL injury risk, particularly during dynamic landing tasks.^1,4-6^ An elevated KAM is characterized by medial knee collapse, increased lateral tibiofemoral joint compression, and internal rotation of the tibia as the shank deviates laterally relative to the femur during weight acceptance.^2^ ACL injury often occurs rapidly after ground contact, coinciding or in close proximity to the occurrence of peak GRF and KAM when the knee is relatively extended, reinforcing the concept that limited knee flexion (i.e., a “stiffer spring”) compromises shock absorption and increases injury risk.^2,4-6^

The quadriceps are a large phasic muscle group that control sagittal knee flexion through eccentric contraction. Strong quadriceps may help to maintain sagittal plane dominance during landing, restricting the hazardous frontal plane compensation manifesting as elevated KAM in athletes whose sport places high demand on the lower extremity.^1-5,7^ As the primary extensors of the knee, it is also reasonable to assume that weaker quadriceps lead to a stiffer landing characterized by limited knee flexion and thus limited force attenuation in the sagittal plane.^2,4,6,8-10^ This shift in loading is clinically relevant, as increased frontal plane kinematics are consistently associated with a higher risk of ACL injury.^4,5,10^ Additionally, quadriceps strength may serve as an indicator of total body strength; thus, athletes lacking strength in this muscle group may be weaker overall, which may contribute to additional risky movement patterns.

The purpose of this study is to examine the relationship between peak KAM and quadriceps strength during landing from a drop jump in adolescent athletes. Based on the neuromuscular structures and landing mechanics involved in shock absorption in the sagittal versus frontal plane, we hypothesize that weaker quadriceps strength will be moderately associated with increased peak KAM.

## METHODS

### Study Design and Participants

The present investigation was an observational secondary analysis of data previously collected as part of the 2024 Baseline ACL Injury Risk Screening and Normative Data study (ClinicalTrials.gov ID: NCT06635668). One hundred and thirty-four healthy adolescent athletes (15.1 ± 1.6 y, 1.70 ± 0.10 m, 66.5 ± 19.0 kg) were originally enrolled in the ACL BUS 2024 cohort. Eligible athletes were free of injuries that limited full activity or sport participation and reported no suspected pregnancy. All participants demonstrated at least a recreational level of physical activity, with many engaged in competitive athletics. The Sanford Health Institutional Review Board gave ethical approval for this work (Study ID: 2976). Written informed consent or assent was obtained from all athletes, and parental consent was secured for minors.

### Data Collection Procedures

Testing was conducted at high schools in the Upper Midwest in conjunction with summer sport and strength and conditioning camps. Athletes completed a warm-up as part of their sport practice or lifting sessions and were then sent in small groups to complete the injury risk screen during the remainder of the session. Athletes had their height, weight, and dominant leg recorded. The dominant leg was identified by asking athletes which leg they would use to kick a soccer ball. The injury risk screen included a test of their quadriceps strength using an isokinetic dynamometer and a test of their movement quality during a drop jump.

To assess muscle strength, athletes were seated in an isokinetic dynamometer (Humac-Norm, CSMi, Stoughton, MA). The administrator positioned and secured each athlete in the dynamometer’s chair and provided a standardized verbal explanation of the test protocol.

Athletes sat fully back in the chair with the lumbar spine in contact with the backrest, which was angled at approximately 85° from horizontal. The backrest position was adjusted so that, at 90° of knee flexion, there was about a 0.5-inch gap between the posterior shank and the front edge of the seat. The seat assembly and dynamometer arm were then translated and rotated to align the estimated knee joint axis with the axis of rotation of the dynamometer. The dynamometer lever arm was positioned so that the pad contacted the distal shank just proximal to the malleoli and was secured with Velcro straps. A thigh strap was placed over the distal anterior thigh of the test leg, while the contralateral leg remained unsupported. Athletes were instructed to maintain contact with the backrest and hold the dynamometer handles throughout the test. The range of motion was standardized from 0° (full extension) to 90° (full flexion), with the anatomical reference angle set at 90°. Prior to testing, athletes performed a trial movement to confirm comfort and absence of mechanical obstruction.

Each leg completed two sets of reciprocal concentric knee extension and flexion at an angular velocity of 60°/s. The first set consisted of five repetitions and served as a familiarization trial, performed at progressively increasing submaximal intensity. After a 20-second rest, the second set consisted of five maximal-effort repetitions through the full range of motion. Administrators delivered standardized verbal instructions and provided synchronized encouragement throughout the test set. The order of limb testing was alternated across athletes to reduce order effects and minimize dynamometer reconfiguration time. Repetitions were required to demonstrate smooth, complete motion and sustained maximal effort in both extension and flexion. If any repetition failed to meet these criteria, the set was repeated following a brief rest. The peak instantaneous concentric knee extension torque for the quadriceps on the dominant limb was computed.

For the drop jump test, athletes began each trial standing upright on a 12-inch (0.30 m) plyometric box positioned 6 inches (0.15 m) from the leading edge of the force plates. At the start signal, athletes stepped forward off the box with both feet simultaneously, landed bilaterally on the force plates, and immediately performed a maximal vertical jump. Athletes were instructed not to lead with one leg to avoid asymmetric landing mechanics and, once they landed, to jump as quickly and as high as possible. No additional coaching or directions were provided. Each athlete performed one practice trial with additional practice provided as needed. Two successful test trials were then collected. Trials were considered unsuccessful if athletes led with a single limb from the box, failed to land entirely on the force plates, contacted both plates with one foot, or stepped off the plates before performing the vertical jump.

Motion data were acquired using a Qualisys Miqus Video motion capture system (Qualisys AB, Gothenburg, Sweden) sampling at 110 Hz and synchronized with ground reaction force data collected from dual side-by-side force plates (4060-07, Bertec Corporation, Columbus, OH, USA) sampling at 880 Hz. Three-dimensional pose estimation was performed using Theia3D (Version 2023.1.0.3160 p18, Theia Markerless Inc, Kingston, ON, Canada) with its inverse kinematics model filtered at a cutoff frequency of 20 Hz. Data were further processed in Visual3D (Version 2024.06.1, HAS-Motion Inc, Kingston, ON, Canada). Analog, as well as kinematic and joint kinetic, data were filtered using a 20 Hz dual-pass Butterworth filter. The peak external KAM on the dominant limb was calculated during the landing phase, defined as the interval between initial contact (vertical ground reaction force >10 N) and toe-off (vertical ground reaction force <10 N), and the average of the two trials was computed.

### Statistical Analysis

The primary variables were peak external KAM during the drop jump landing phase on the dominant leg and peak isokinetic concentric knee extension torque, or quadriceps strength, on the dominant leg. Both quadriceps strength and KAM values were normalized to body mass to account for inter-subject variability. A simple linear regression model was used to examine the association between normalized quadriceps strength and peak external KAM. Model residuals were evaluated for normality by inspection of Q–Q plots and significance was at p < 0.05.

To address the stated research question and hypothesis, an a priori power analysis was conducted to detect a moderate effect size (f^2^ = 0.15) with significance level of 0.05 and power of 0.80, which revealed a minimum of 54 athletes would be required.

## RESULTS

The linear regression model revealed that although there was a negative trend suggesting that greater quadriceps strength may be associated with slightly lower KAM (−.053 (95% CI [−.137, .030])), the relationship was not statistically significant (F(1,119) = 1.618, R^2^ = 0.013, f^2^ = 0.013, p = 0.206) (Figure 1).

**Figure 1.**
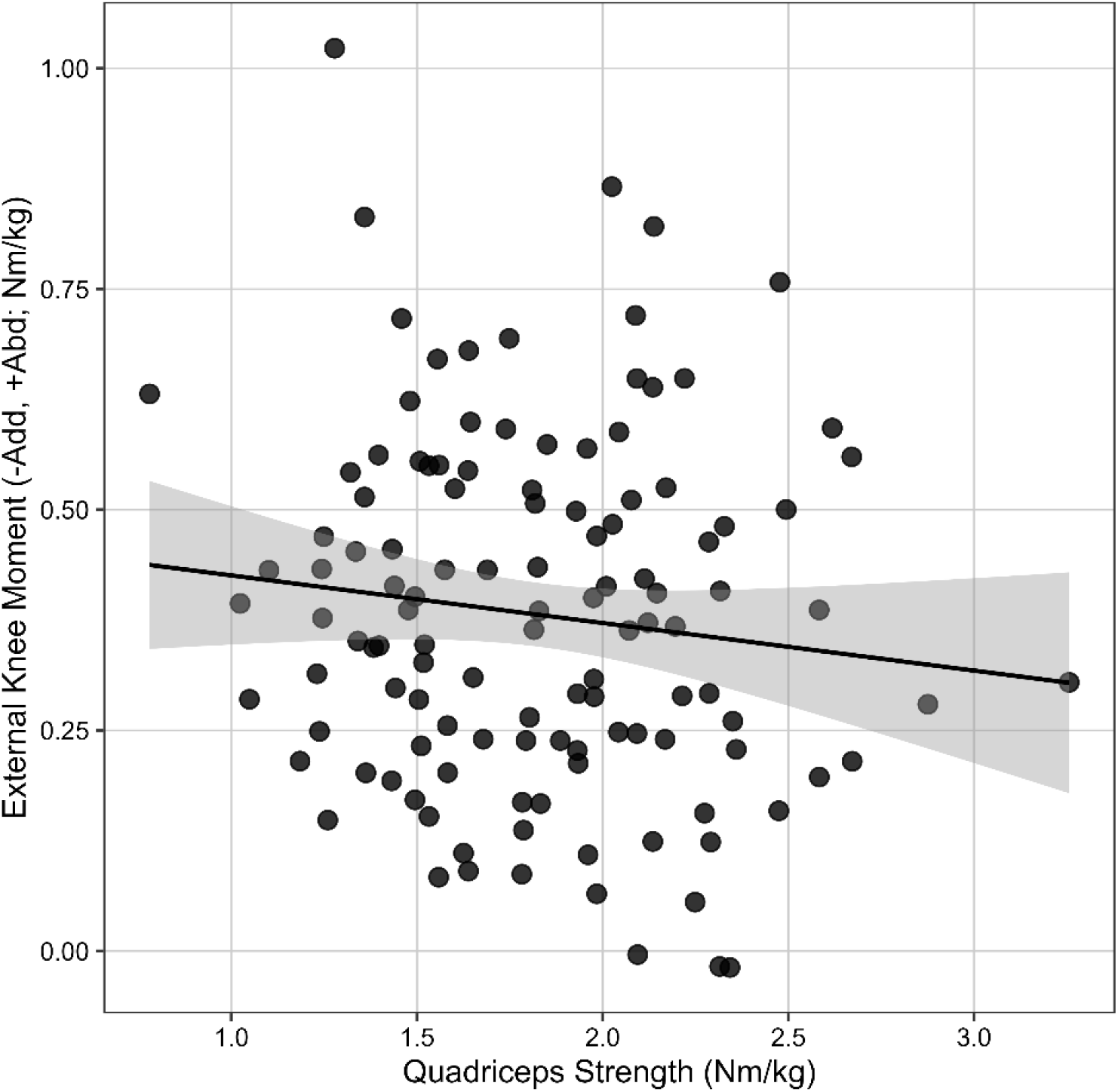
Linear regression between peak knee extension moment (quadriceps strength) normalized to body mass (Nm/kg) and peak external frontal plane knee moment (-Adduction/+Abduction) normalized to body mass (Nm/kg). Black line: linear regression; grey ribbon: 95% confidence interval; black dots: individual athletes.

## DISCUSSION

The purpose of this study was to examine the relationship between peak KAM, a known biomechanical risk factor for ACL injury, and quadriceps strength during landing from a drop jump in adolescent athletes. Based on the neuromuscular structures and landing mechanics involved in shock absorption in the sagittal versus frontal plane, we hypothesized that weaker quadriceps strength would be moderately associated with increased peak KAM. No statistically significant association between these variables was identified in this study; therefore, the hypothesis was not supported.

Prior literature provides important context for the present findings. Foundational work by Hewett et al. established frontal-plane knee loading, quantified by KAM, as a strong predictor of ACL injury risk.^1^ Work by Pollard et al. established the sagittal-to-frontal plane relationship, finding that a high flexion landing pattern was associated with reduced frontal plane loading, thus decreasing strain on the ligaments of the knee.^9^ Hewett also demonstrated that sagittal-plane measures influenced by the quadriceps are not associated with ACL injury risk,^1^ and Pollard found that landing with higher knee flexion was associated with decreased vastus lateralis activation,^9^ findings that may help explain the absence of an association observed in the present study. Sigurđsson et al. found that greater knee flexion excursion—which requires a larger internal knee extension moment and thus places a greater demand on the quadriceps—was associated with an increased frequency of large early peak KAM.^2^ Similarly, Huang et al. found that athletes with lower quadriceps strength capacity, as measured by rate of torque development during a separate strength assessment, tended to exhibit less knee flexion during a single-leg stop-jump task, a landing pattern associated with elevated ACL injury risk.^6^ Greater quadriceps strength capacity appears to be associated with greater knee flexion during landing, which increases internal knee extension moment demand and may allow athletes to adopt deeper, more sagittal-plane–dominant landing strategies. Greater knee flexion, in turn, is associated with lower KAM.

KAM reflects frontal-plane mechanics, occurring most when the body must control side-to-side motion, whereas the quadriceps primarily regulate sagittal-plane motion about the knee, making the absence of an association between the two biomechanically logical. If quadriceps strength does influence KAM, assessing athletes while performing a lateral or cutting-based movement rather than a vertical jump may better elicit a KAM and demonstrate an association. Although KAM has been described as a compensatory strategy that may emerge when sagittal-plane attenuation—controlled largely by the phasic quadriceps—is insufficient, the present findings suggest that frontal-plane compensation is driven more so by overall movement strategy rather than by the maximum strength of the quadriceps alone.^9^ KAM is influenced by hip and trunk positioning as well as neuromuscular coordination; for example, greater engagement of the hip extensors during landing has been shown to reduce frontal-plane knee loading,^9^ while altered trunk positioning can shift the center of mass, increase ground reaction forces, and increase knee ligament strain.^3,5^ Collectively, these findings indicate that KAM arises from coordinated, multi-joint movement patterns rather than the strength of a single muscle group, providing a likely explanation for the absence of an association in the present study. This underscores the importance of neuromuscular, whole-body approaches to ACL injury risk reduction.

A key limitation of this study is that KAM and quadriceps strength were assessed using different tasks, with KAM measured during a dynamic landing task and quadriceps strength measured isokinetically. Quadriceps strength during landing is eccentric, working rapidly to slow the knee down, but athletes in the present study were tested concentrically. These discrepancies in assessment modalities and movements tested may have limited our ability to detect meaningful associations between these variables. There are many ways to measure strength, so future studies could address this limitation by employing methods to assess quadriceps strength to more closely reflect neuromuscular activation during sport-specific movements and thus better quantify the relationship between movement and strength. Isometric testing would better reflect an athlete’s ability to rapidly develop force, which may be more important for controlling knee motion right after the foot hits the ground and peak KAM arises. This idea assumes that how fast the quadriceps can generate a force generally reflects how fast other muscle groups, like the hip abductors and trunk muscles, can respond to promote knee alignment. Additionally, maximal quadriceps strength may not accurately represent the neuromuscular activation patterns of this muscle group during the drop jump task from which KAM was calculated, or during athletic maneuvers in which ACL injuries occur secondary to high early peak KAM. How strength is measured thus may matter more than how strong someone is. Incorporation of electromyography (EMG) to evaluate muscle activation timing and co-contraction patterns during landing tasks may further inform the relationship between quadriceps function and KAM. Finally, although standardized protocols were used to assess quadriceps strength and the drop jump, involvement of multiple testers may have introduced inter-tester variability. This potential source of error can be reduced through further standardization in future studies.

The absence of a direct association between quadriceps strength and KAM in the present study is a meaningful finding that refines the current understanding of ACL injury biomechanics. The primary aim was to improve understanding of how landing during tasks such as a drop jump may lead to frontal-plane compensatory strategies in the presence of insufficient sagittal-plane energy attenuation. The findings of the present study suggest that quadriceps strength alone may be insufficient to explain variations in frontal-plane knee loading manifested as KAM. Further research should investigate the roles of additional muscle groups and neuromuscular coordination in modulating KAM, with the goal of informing preventive intervention strategies to reduce the burden of ACL injury. The assessment of individual athletes’ movement patterns through accessible, context-specific data collection—as demonstrated in this study—may facilitate personalized interventions targeting modifiable risk factors like KAM, ultimately supporting the short- and long-term musculoskeletal health and performance of athletes.

## Data Availability

Data produced in the present study are available upon reasonable request to the authors.

## REFERENCES

1. Hewett TE, Myer GD, Ford KR, et al. Biomechanical measures of neuromuscular control and valgus loading of the knee predict anterior cruciate ligament injury risk in female athletes: a prospective study. Am J Sports Med. 2005;33(4):492–501. doi:10.1177/0363546504269591

2. Sigurđsson HB, Karlsson J, Snyder-Mackler L, Briem K. Kinematics observed during ACL injury are associated with large early peak knee abduction moments during a change of direction task in healthy adolescents. J Orthop Res. 2021;39(10):2281–2290. doi:10.1002/jor.24942

3. Hewett TE, Torg JS, Boden BP. Video analysis of trunk and knee motion during non-contact anterior cruciate ligament injury in female athletes: lateral trunk and knee abduction motion are combined components of the injury mechanism. Br J Sports Med. 2009;43(6):417–422. doi:10.1136/bjsm.2009.059162

4. Ward SH, Blackburn JT, Padua DA, et al. Quadriceps Neuromuscular Function and Jump-Landing Sagittal-Plane Knee Biomechanics After Anterior Cruciate Ligament Reconstruction. J Athl Train. 2018;53(2):135–143. doi:10.4085/1062-6050-306-16

5. Tsatalas T, Karampina E, Mina MA, et al. Altered Drop Jump Landing Biomechanics Following Eccentric Exercise-Induced Muscle Damage. Sports (Basel). 2021;9(2):24. Published 2021 Feb 5. doi:10.3390/sports9020024

6. Huang YL, Chang E, Johnson ST, Pollard CD, Hoffman MA, Norcross MF. Explosive Quadriceps Strength and Landing Mechanics in Females with and without Anterior Cruciate Ligament Reconstruction. Int J Environ Res Public Health. 2020;17(20):7431. Published 2020 Oct 13. doi:10.3390/ijerph17207431

7. Kernozek TW, Torry MR, VAN Hoof H, Cowley H, Tanner S. Gender differences in frontal and sagittal plane biomechanics during drop landings. Med Sci Sports Exerc. 2005;37(6):1003–1013. doi: 10.1249/01.mss.0000171616.14640.2b

8. Hart JM, Ko JW, Konold T, Pietrosimone B. Sagittal plane knee joint moments following anterior cruciate ligament injury and reconstruction: a systematic review [published correction appears in Clin Biomech (Bristol, Avon). 2010 Aug;25(7):749. Pietrosimione, Brian [corrected to Pietrosimone, Brian]]. Clin Biomech (Bristol). 2010;25(4):277–283. doi:10.1016/j.clinbiomech.2009.12.004

9. Pollard CD, Sigward SM, Powers CM. Limited hip and knee flexion during landing is associated with increased frontal plane knee motion and moments. Clin Biomech (Bristol). 2010;25(2):142–146. doi:10.1016/j.clinbiomech.2009.10.005

10. Shultz SJ, Pye ML, Montgomery MM, Schmitz RJ. Associations between lower extremity muscle mass and multiplanar knee laxity and stiffness: a potential explanation for sex differences in frontal and transverse plane knee laxity. Am J Sports Med. 2012;40(12):2836–2844. doi:10.1177/0363546512461744

